# Identification of disease-specific extracellular vesicle-associated plasma protein biomarkers for Duchenne Muscular Dystrophy and Facioscapulohumeral Muscular Dystrophy

**DOI:** 10.1101/2024.11.29.24317861

**Authors:** Mustafa Bilal Bayazit, Don Henderson, Kim Truc Nguyen, Eduardo Reátegui, Rabi Tawil, Kevin M. Flanigan, Scott Q. Harper, Nizar Y. Saad

**Author notes:** Corresponding author: Nizar Y. Saad, PhD. Corresponding author’s address: Jerry R. Mendell Center for Gene Therapy, Abigail Wexner Research Institute at Nationwide Children’s Hospital, 575 Children’s Crossroad Columbus, OH 43215, USA. Corresponding author’s. Corresponding author’s.

## Abstract

**Objective:** Reliable, circulating biomarkers for Duchenne, Becker and facioscapulohumeral muscular dystrophies (DBMD and FSHD) remain unvalidated. Here, we investigated the plasma extracellular vesicle (EV) proteome to identify disease-specific biomarkers that could accelerate therapy approvals.

**Methods:** We extracted EVs from the plasma of DBMD and FSHD patients and healthy controls using size-exclusion chromatography, conducted mass spectrometry on the extracted EV proteins, and performed comparative analysis to identify disease-specific biomarkers. We correlated the levels of these biomarkers with clinical outcome measures and confounding factors.

**Results:** The muscle-associated proteins PYGM, MYOM3, FLNC, MYH2 and TTN were exclusively present in DBMD EVs. PYGM, MYOM3, and TTN negatively correlated with age. PYGM and MYOM3 levels were elevated in patients without cardiomyopathy, and PYGM levels were specifically elevated in ambulatory DMD patients. On the other hand, female FSHD patients displayed significantly higher MBL2 and lower GPLD1 levels. However, male FSHD patients exhibited higher C9 and lower C4BPB levels. Additionally, desmosome proteins JUP and DSP were uniquely found in FSHD males. MBL2 positively correlated with age and C4BPB negatively correlated with FSHD severity in male patients.

**Interpretation:** Our findings underscore the sensitivity of analyzing circulating EV content to identify disease-specific protein biomarkers for DBMD and FSHD. Our results also emphasize the potential of EV-based biomarker discovery as a promising approach to monitor disease progression as well as effectiveness of therapies in muscular dystrophy, potentially contributing to their approval. Further research with larger cohorts is needed to validate these biomarkers and explore their clinical implications.

## Introduction

Duchenne Muscular Dystrophy (DMD) is a severe X-linked neuromuscular disorder affecting approximately 1 in 3,500 to 1 in 5,200 boys, making it the most common form of muscular dystrophy [1–3]. DMD is characterized by progressive muscle degeneration and atrophy. Patients typically lose ambulation by the second decade, with death often occurring in the third decade of life. The underlying pathophysiology involves mutations in the *DMD* gene, leading to a complete absence of dystrophin, a critical cytoskeletal protein essential for maintaining myofiber integrity. Becker Muscular Dystrophy (BMD) is a milder allelic variant caused by mutations in the same gene. In contrast to DMD, BMD patients retain some dystrophin function, leading to a more variable clinical course and later disease onset [1, 4]. On the other hand, facioscapulohumeral muscular dystrophy (FSHD) is the third most common muscular dystrophy affecting 1 in 8,333 people. It causes progressive skeletal muscle weakness and wasting. FSHD is an autosomal dominant disease caused by de-repression of the transcription factor DUX4 in skeletal muscles, leading to cell death, oxidative stress, and other muscle abnormalities [5].

Most DBMD patients rely on corticosteroid treatments, other oligonucleotide therapies and the recently approved microdystrophin gene replacement therapy to slow disease progression and restore dystrophin expression, with limited clinical benefit [6–9]. Nevertheless, other potentially more effective therapies have entered clinical trials [10–12]. By contrast, FSHD patients still lack any approved, effective therapy, although few treatments are currently in or about to enter clinical trials [13]. While the recent developments in DBMD and FSHD treatments are encouraging, identifying clinical outcome measures to validate these therapies remains an ongoing effort in the field. Functional, imaging, biopsy-based and patient-reported outcome measures have been used to evaluate therapeutic efficacy in DBMD and FSHD clinical trials [8]. Additionally, patient-reported outcomes (PROs), which capture the patient’s perspective on their quality of life (QoL) and disease impact are used. Despite their rigorous validation, these clinical outcome measures remain labor intensive, invasive, time consuming, inconvenient for some patients, and expensive, which could delay approval of therapies. Moreover, biopsy-based biomarkers are unreliable in predicting disease progression and therapeutic efficacy. In the case of Duchenne, assessing dystrophin levels in affected skeletal muscles is not only inconvenient but also unreliable, as dystrophin levels do not consistently correlate with disease severity. Additionally, the natural history of DMD poses challenges for clinical trial design and efficacy assessment [1, 4, 14]. In fact, DMD shows variability in presentation and progression due to factors such as low residual dystrophin expression, environmental influences like steroid use, and genetic modifiers that impact muscle inflammation and fibrofatty infiltration [15, 16]. In the case of BMD, the variability in disease presentation and progression is even greater, depending on the specific *DMD* gene mutation. Similar challenges exist in FSHD. The levels of DUX4 and DUX4-target genes in patient muscle biopsies were not significantly reduced when measured in an FSHD clinical trial that tested the therapeutic effects of the DUX4-lowering drug losmapimod [17–20]. This was most probably due to the rare presence of DUX4 in only 0.01 to 0.1% of patient myonuclei and to the unpredictability of FSHD progression and challenges in selecting muscle biopsies that express or previously expressed DUX4 [21]. Consequently, the existence of reliable minimally invasive circulating biomarkers to assess therapeutic efficacy would be a valuable addition to the existing functional outcome measures. Such biomarkers would allow a better stratification of patients for clinical trials, better tracking of disease progression and detection of early signs of response to therapy that would anticipate clinical benefits. Additionally, they would allow longitudinal monitoring of treated patients to assess persistence of therapeutic interventions.

So far, creatine kinase isoforms (CK-MB and CK-MM) are the most widely used circulating biomarkers for DMD. These CK isoforms are nonspecific indicators of muscular dystrophy and inflammation, but do not correlate with disease progression. Moreover, age, residual muscle mass and physical activity are all factors that influence CK levels [22]. Other muscle-related serum proteins such as alanine aminotransferase (ALT), aspartate aminotransferase (AST), lactate dehydrogenase (LDH), and alkaline phosphatase (ALP) showed promising results in pre-clinical studies, but their clinical relevance remains to be confirmed [22]. Likewise, only subtle changes in plasma complement components, indicators of general innate immunity, were observed in FSHD patients [23]. On the other hand, other circulating biomarkers identified in FSHD still require validation in larger cohorts [24, 25]. Altogether, there is still a critical need to discover reliable and minimally invasive molecular biomarkers for DBMD and FSHD.

Extracellular vesicles (EVs) are non-replicative, lipid-bilayer delimited particles that are naturally released from most cells. EVs carry tissue-specific molecules (e.g., lipids, nucleic acids and proteins). Secreted EVs can either be captured by neighboring cells or released into biofluids (e.g., blood), functioning as messengers in cell-to-cell or inter-tissue communication. Importantly, circulating (blood plasma) EVs are emerging as important disease biomarkers as their molecular content can inform the pathophysiological state of the disease [26]. Noticeably, EVs derived from patient plasma are generally more enriched for pathogenic proteins and nucleic acids than crude plasma [27]. The active packaging of some proteins and nucleic acids into EVs led us to investigate their contents as potential disease-specific circulating biomarkers for DBMD and FSHD. Our hypothesis is that the selective packaging of EV content is likely influenced by the disease-related mechanisms driving muscle degeneration, making them reflective of these underlying pathological processes. Interestingly, despite the significant contribution of muscles to the circulating EV pool, EV-protein content of a biofluid has not yet been extensively analyzed in muscular dystrophy patients [28–30]. Additionally, no previous work has conducted a comprehensive, unbiased proteomic profiling of circulating DBMD and FSHD EVs similar to our approach.

In this paper, we unraveled the proteomic profile of circulating EVs in DBMD and FSHD patients, and identified prospective disease-specific protein biomarkers. Our correlation analyses revealed that some biomarkers were associated with both disease phenotypes and confounding variables.

## Subjects/Materials and Methods

### Patient selection

De-identified plasma samples for the FSHD cohort were obtained from Dr. Rabi Tawil (University of Rochester, NY, USA) as part of the clinical trial readiness to solve barriers to drug development in FSHD” (ReSolve) study [31]. In the FSHD cohort, clinical outcome measures included the clinical severity score (CSS), the FSHD composite score (FSHD-COM), and D4Z4 contraction size. The CSS is a 10-grade system that scores the extent of weakness in various body regions, where a score of 0 indicates no muscle weakness and 10 indicates wheelchair dependency [32]. The FSHD-COM score is a comprehensive, 18 item performance-based functional composite outcome measure assessing the legs, shoulders and arms, trunk, hands, and balance/mobility [33]. De-identified human plasma samples of dystrophinopathy and normal control subjects were obtained through the neuromuscular clinic at Nationwide Children’s Hospital following informed consent obtained under a protocol approved by the Institutional Review Board at Nationwide Children’s Hospital (Columbus, OH, USA) (**Table 1**). For all samples, plasma was collected using anti-coagulant (EDTA) treated collection tubes, and each sample was assigned a blinding code so that all batches of samples were run in a blinded fashion.

**Table 1.**
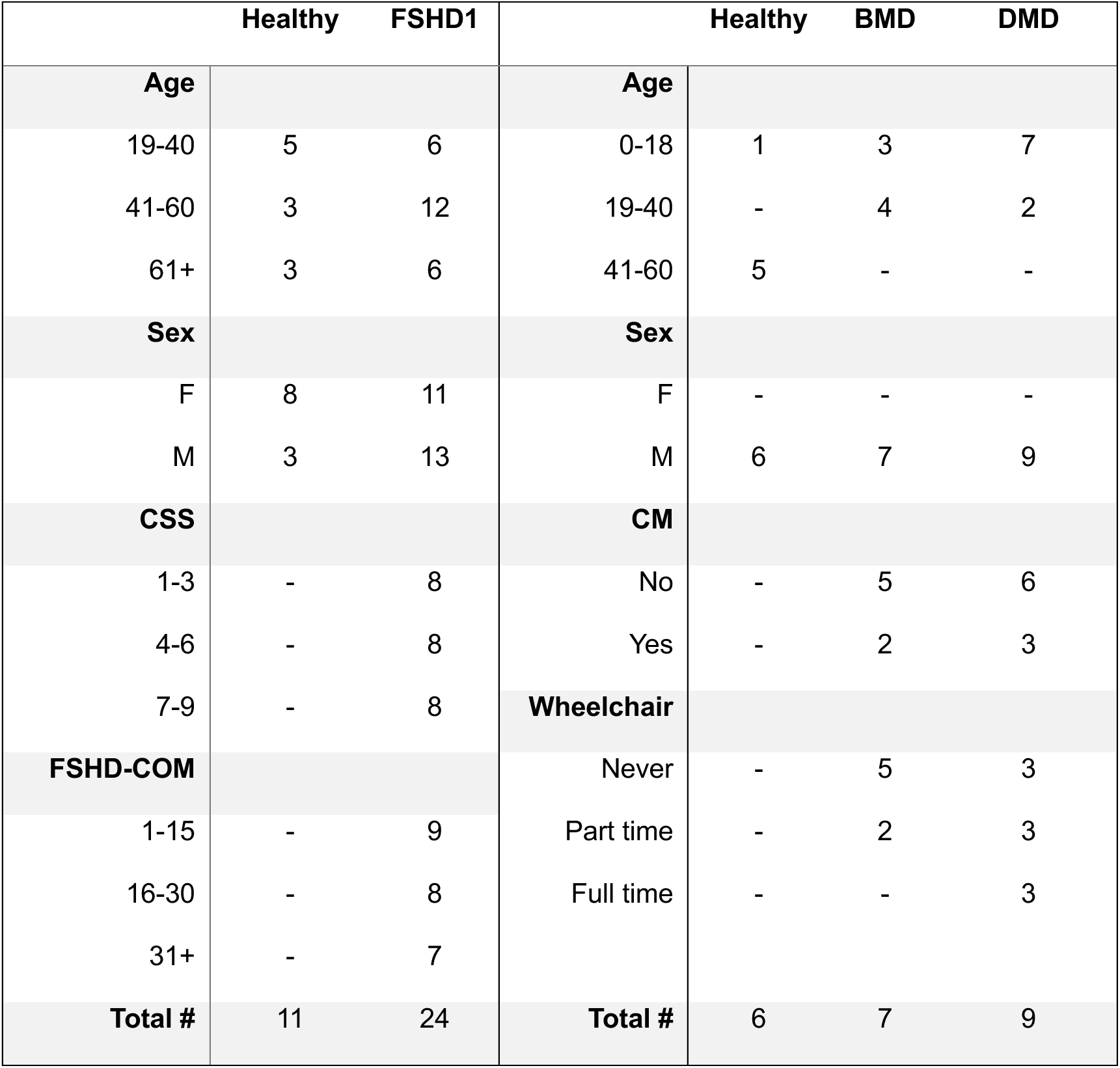
Patient clinical data of two cohorts. CSS: Clinical Severity Score. FSHD-COM: FSHD composite score. CM: Cardiomyopathy.

### EV isolation

For proteomic analyses, plasma EVs were isolated by size-exclusion chromatography (SEC), following the most recent guidelines from the International Society for Extracellular Vesicles for isolation, storage, and characterizations [34]. This method allows for efficient EV separation from soluble components and most lipoproteins [35]. While there are various other ways to isolate EVs (e.g., filtration, differential centrifugation, density gradient and immunoprecipitation), most of these methods are exclusively targeted for either EV yield or specificity. In contrast, SEC is efficient in both parameters, and therefore ideal for biomarker discovery [36]. 500 µL pre-cleared plasma samples were loaded onto Izon qEV original chromatography columns (35 nm) and using an automated fraction collector (Izon), the first five fractions were collected and combined. EV size and concentration were monitored by Nanoparticle Tracking Analysis (NTA, Nanosight) with a detection threshold of three. EV structure was further characterized by transmission electron microscopy (TEM). EV quality was assessed by western blot to evaluate the presence of plasma EV markers (e.g., CD9) and plasma proteins (e.g., ApoB, C3). To eliminate bias in subsequent analyses, EV isolation was performed blinded. Samples were then batched to ensure patient and healthy controls samples were analyzed together.

### Protein isolation and Mass spectrometry

The plasma EVs, isolated by SEC, were lysed with 5% SDS and sonicated to extract proteins. A total of 100 μg of protein was used for mass spectrometry (MS), while the rest was stored for western blot analysis. For MS, the proteins were digested via Strap or Rapigest depending on the sample quantity and following the Core’s protocol. Liquid Chromatography was then performed with tandem MS (LC-MS/MS). Using Mascot Daemon by Matrix Science version 2.7.0 (Boston, MA) via Proteome Discoverer (version 2.4 Thermo Scientific), our data was searched against the most recent Uniprot databases. Label free quantitation was performed using the spectral count approach, in which the relative protein quantitation is measured by comparing the number of MS/MS spectra identified from the same protein in each of the multiple LC-MS/MS datasets.

### Statistical Analysis

Differential expression of both EV-associated protein levels was assessed using the Wald test with a significance cut-off of *p* <0.05 (R package: DESeq2). To control for testing of many proteins, false discovery rates (FDR) were computed from raw *p* values, with FDR <0.1 and log2FC >1 used as a cutoff for significance. Pearson correlation coefficients with *p* values were computed on scatter plots with the “stat_cor” function in R.

### Immunoblotting

Select biomarkers identified by mass spectrometry were verified with western blot. To verify the EV association of the biomarkers (i.e., inside or outside EVs), EVs were treated with Proteinase K (20 µg/ml) for 1 hour (h) at 37°C, to only retain the proteins packed within the EVs and protected by their lipid bilayer membrane. PMSF (20 µM) was added to the EVs 1h at room temperature (RT) post treatment to inhibit Proteinase K and ensure it would not be accessible to EV proteins prior to western blot. As additional control, EV membranes were treated simultaneously with Proteinase K and 1% Triton X-100, which ruptures EV membrane, allowing Proteinase K to digest intraluminal EV proteins. EVs were then boiled in Laemmli buffer and proteins were size-separated using a 4-12% Bis-Tris or 3-8% Tris-Acetate denaturing gels (NuPAGE, Invitrogen). The proteins were transferred to a PVDF membrane and blocked with 5% milk in TBS-T for 1h at RT. The primary antibodies were diluted in the blocking buffer and incubated overnight at 4°C. After washing off the primary antibodies with TBS-T, membranes were incubated with an HRP-conjugated secondary antibody against the host species of the primary antibody for 1h at RT. Primary antibodies used in this study were: rabbit anti-CD9 (20597-1-AP, 1:4000, Proteintech), rabbit anti-MBL2 (24207-1-AP, 1:1000, Proteintech), rabbit anti-MYOM3 (17692-1-AP, 1:1000, Proteintech), rabbit anti-PYGM (19716-1-AP, 1:1000, Proteintech), mouse anti-γ-catenin (JUP) (sc-514115, 1:500, Santa Cruz Biotechnology), rabbit anti-C3 (21337-1-AP, 1:10000, Proteintech), and rabbit anti-ApoB (20578-1-AP, 1:1500, Proteintech). The membranes were stripped using the Restore Western Blot Stripping Buffer (21059, Thermo Fisher) for 30 mins at room temperature. As protein molecular weight marker, the precision plus protein standards dual color from Bio-Rad (161-0374) was used. All blots were developed using the ChemiDoc MP Imaging System (Bio-Rad).

## Results

### Isolation and characterization of plasma EVs

Analysis was performed in two cohorts. The first consisted of FSHD patients compared to normal controls, and the second consisted of dystrophinopathy patients compared to normal controls (**Table 1**).

Under TEM, EVs displayed a nanoparticle circular shape delimited by a bilipid membrane (**Fig. 1A**). Our characterization of the size and concentration of EVs, revealed that DMD patients had 1.10-fold larger EVs than BMD patients (108.6 nm vs 98.30 nm, N=7-9, *p*val: 0.045, unpaired t-test), FSHD patients had 1.3-fold smaller EVs than healthy controls (137.8 nm vs 109 nm, N=5, *p*val: 0.04, unpaired t-test). Plasma EV concentration also significantly varied between the groups. In BMD patients, EV concentration was 25-fold lower than healthy controls (1.1×10^11^ particles/mL vs 2.8×10^12^ particles/mL, N=7-9, *p*val: 0.01, unpaired t-test). Similarly, EV concentration was 2.3-fold lower in FSHD patients than in healthy controls (0.9×10^12^ particles/mL vs 2.1×10^12^ particles/mL, N=5, *pval*: 0.01, unpaired t-test) (**Fig. 1B-E**).

**Figure 1.**
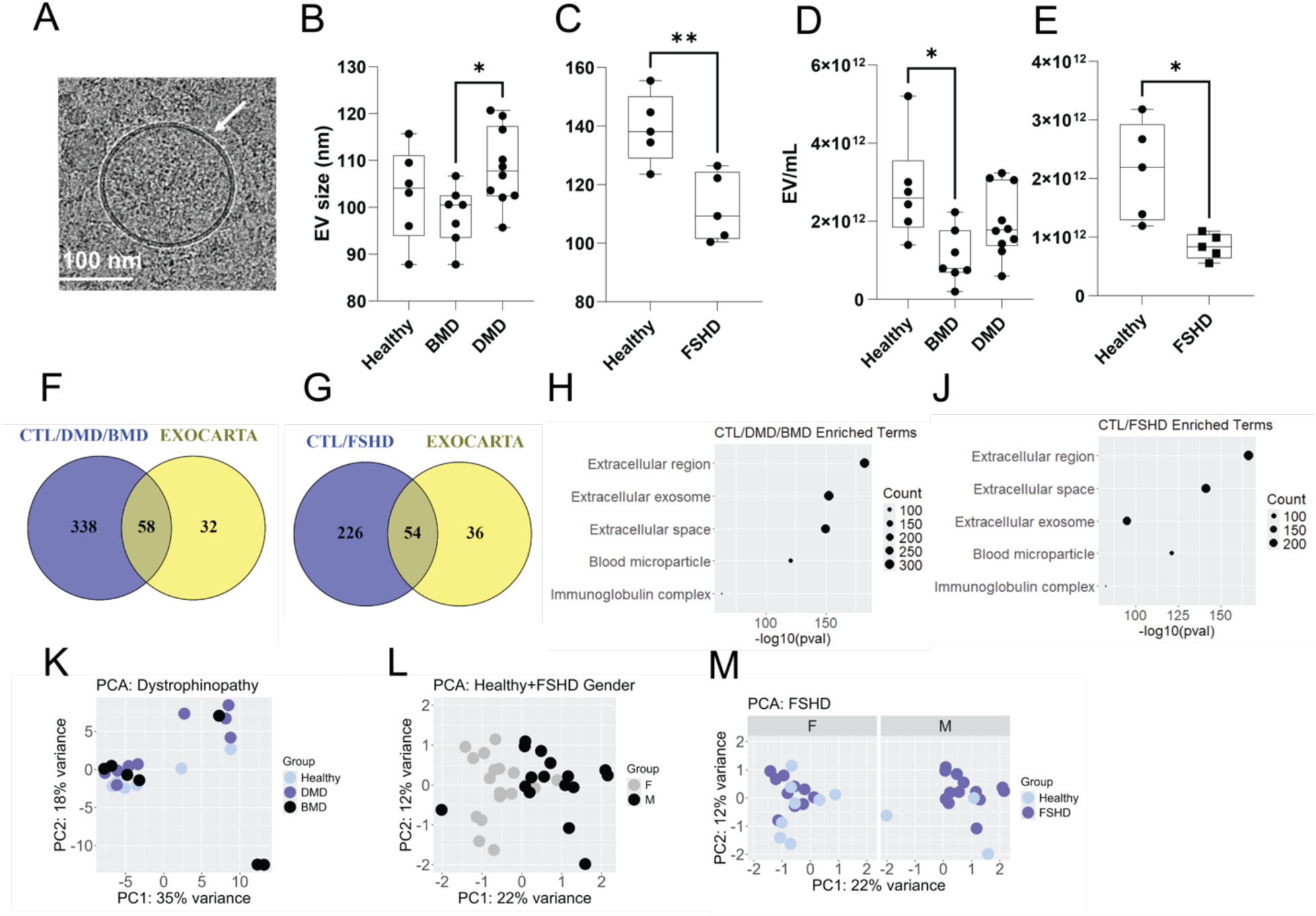
Characterization of plasma EVs of Dystrophinopathy and FSHD1 patients and healthy controls by Size Exclusion Chromatography (SEC). **(A)** Transmission electron microscopy (TEM) image showing a control EV and its lipid bilayer membrane. **(B-E)** Size and concentration of EVs determined by Nanoparticle Tracking Analysis (NTA). **(F and G)** Venn diagram of overlapping proteins between this study and the Exocarta Database for plasma EV proteins. **(H and J)** Enrichment analysis of proteins identified in this study. Enrichment analysis is conducted with DAVID software for Gene Ontology (GO) Terms with Cellular Compartment sub-analysis. Top five significantly (Bonferroni corrected *p*val<0.05) enriched terms are displayed. **(K-M)** Principal component analysis (PCA) of proteins identified by mass spectrometry. FSHD cohort sub-grouped according to sex **(M)** ∗ *p*val < 0.05, ∗∗ *p*val < 0.01 by two-tailed unpaired t-test (BMD, N=7; DMD, N=9; cognate healthy controls: N=6; FSHD1, N=24; cognate healthy controls, N=11).

Following EV characterization, we set out to reveal their protein content by performing mass spectrometry (LC-MS/MS) analysis. We identified 396 and 280 proteins in dystrophinopathy and FSHD1 cohorts, respectively, with a mean spectral count per condition greater than 1. Proteins indexed in ExoCarta EV plasma database (http://exocarta.org, accessed on July 23, 2024) were used as reference (**Fig. 1F-G**). We then conducted the Gene Ontology (GO) enrichment analysis using the online DAVID tool with Cellular Compartment (CC) sub-analysis [37]. This illustrated that the analyzed EVs are enriched in ‘Extracellular region’, ‘Extracellular exosome’, and ‘Extracellular space’ proteins (**Fig. 1H-J**). To unveil any potential condition-specific clustering of overall protein content, we ran principal component analyses (PCA). As a result, we did not detect any specific clustering of samples in DBMD patients (**Fig. 1K**). While dystrophinopathy only affects males, our FSHD1 cohort included both male and female patients. In line with reports of sex specific differences in circulating EV content [38], we observed plasma EV content to cluster based on sex (**Fig. 1L**). Therefore, we subsequently analyzed the female and male FSHD1 cohorts, independently. Like the dystrophinopathy cohort, no disease specific clustering of samples was observed in FSHD patients (**Fig. 1M**).

### Disease-specific differences in plasma EV proteomes

To reveal any potential plasma EV biomarkers, we ran differential protein expression analyses between patient samples and their corresponding healthy controls. Using stringent cut-off criteria (FDR adjusted *p*.value [*p*.adj]<0.1, *p*val<0.05 and log2FC >1 | log2FC <-1), we identified proteins dysregulated in each disease (**Fig. 2**). In DBMD cohorts formed of 7 BMD, 9 DMD and 6 healthy controls, all proteins that met the cut-off criteria were found to be exclusively expressed in patient EVs. These dystrophinopathy-exclusive proteins are Myomesin 3 (MYOM3), Glycogen phosphorylase, muscle associated (PYGM), Titin (TTN), and Filamin C (FLNC) (**Fig. 2 A-B)**. Additionally, DMD plasma EVs were also enriched for Myosin heavy chain (MYH2) (**Fig. 2B**). Notably, these dystrophinopathy-specific EV proteins are involved in key muscle functions such as muscle contraction and sarcomere organization, as illustrated by GO enrichment analyses (**Fig. 2C**). In contrast to dystrophinopathy patients, plasma EVs of FSHD1 patients displayed a distinctive profile of differentially expressed proteins, with an overall alteration in the complement pathway components of innate immunity (**Fig. S1**). The female cohort (11 FSHD, and 8 healthy control) displayed the enrichment of Mannose binding lectin 2 (MBL2) (log2FC: 2.20, *p*val: 0.0013, *p*.adj: 0.0722), and the decrease of Glycosylphosphatidylinositol-specific phospholipase D (GPLD1) (log2FC: −1.20, *p*val: 0.0002, *p*.adj: 0.025) in FSHD EVs (**Fig. 2D**). In the male cohort (13 FSHD, 3 healthy control), Junctional Plakoglobin (JUP) and Desmoplakin (DSP) were exclusively expressed in FSHD patients. Moreover, complement component 9 (C9) was increased (log2FC: 1.44, *p*val: 0.0031, *p*.adj: 0.0973), and C4b-binding protein (C4BPB) was decreased (log2FC: −1.02, *p*val: 0.0007, *p*.adj: 0.0527) in male FSHD patients (**Fig. 2E**).

**Figure 2.**
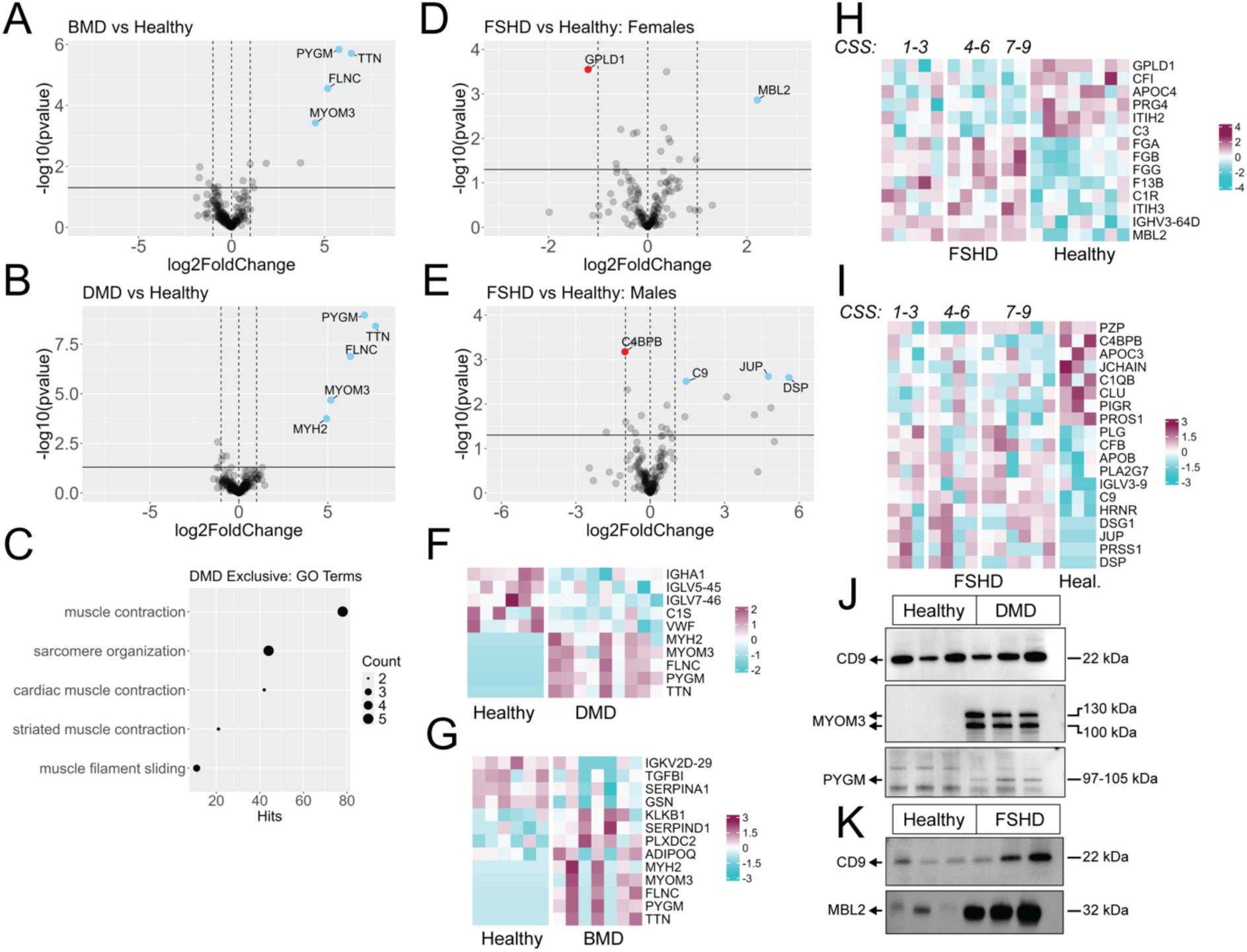
Differential protein levels in plasma EVs of dystrophinopathy and FSHD1 patients and healthy controls. (**A-B**) Volcano plots of the dystrophinopathy cohorts. (**C**) Enrichment analysis on the DMD cohort conducted with DAVID software for Gene Ontology (GO) Terms with Biological Processes as sub-analysis. Top five significantly enriched terms are displayed (Bonferroni corrected pval<0.05). (**D-E**) Volcano plots of the FSHD1 cohorts. For all cohorts, FDR<0.1, pval<0.05 and log2FC >1 (dashed lines) was used as a cutoff for significance. Labeled are proteins that meet the cutoff criteria. **(F-I-)** Corresponding heatmaps of relative levels (i.e., z-scores) displaying all proteins that are significantly changing using nominal *p*val<0.05. For the FSHD cohorts, disease severity based on clinical severity score (CSS) is displayed. **(J-K)** Confirmation of the mass spectrometry results by western blot for MYOM3, PYGM and MBL2 (uncropped blots are in **Supplementary fig. S4**).

While MBL2, C9, and C4BPB are components of the complement pathways of the innate immunity [39], GPLD1 is a circulating membrane regulator that is actively involved in energy metabolism [40], and JUP and DSP are structural desmosome proteins [41]. The diversity and disease specificity of the proteomic changes is further emphasized using heatmaps displaying the relative levels (as indicated by z-score) of all dysregulated protein changes (nominal *p*val<0.05) (**Fig. 2F-I**). Moreover, we validated the major changes observed by mass spectrometry with immunoblotting for MYOM3, PYGM, and MBL2 (**Fig. 2 J-K**). In the DMD cohort, the specific PYGM band at 97-105 kDa can only be observed in patient samples, meanwhile MYOM3 staining yielded in two distinct fragments in patients, which is in line with previous reports on MYOM3 fragmentation [42] (**Fig. 2J**). While not exclusively expressed in patient EVs, the significant enrichment of MBL2 can be observed in female FSHD patients (**Fig. 2K**).

### Correlations of plasma EV proteins with confounding factors

Considering the dynamic and variable nature of dystrophinopathies and FSHD, we set out to correlate the differentially expressed protein levels with confounding factors. For dystrophinopathy patients assessed confounding factors included age, ambulation status, steroid use, and presence of cardiomyopathy. For FSHD, we correlated our findings with age, the FSHD-COM and CSS scores as well as the D4Z4 contraction size. We used Pearson’s correlation analysis to determine the strength of the linear relationship between two variables (*R*), and to determine the statistical significance of the linear relationship (*p*val). Among tested variables, patient age was a major factor determining PYGM, MYOM3, and TTN levels in dystrophinopathy patients (**Fig. 3A-B**). In these cases, we observed significant negative correlations with age (PYGM *R*=-0.74, *p*val: 0.001; MYOM3 *R*=-0.6, *p*val: 0.014; TTN *R*=-0.5, *p*val: 0.049). Furthermore, using one-way ANOVA analyses, we found that both PYGM and MYOM3 levels were significantly higher in patients with no reports of cardiomyopathy (PYGM FC: 3.7, *p*val: 0.03, MYOM3 FC: 4.3, *p*val: 0.02) (**Fig. 3C-D**). PYGM levels were also significantly lower (FC: 0.18, *p*val: 0.02) in patients who are dependent on full-time wheelchair use (**Fig. 3E**). Altogether, it can be speculated that plasma EV-associated PYGM peaks early in DMD pathogenesis, and declines as the disease progresses.

**Figure 3.**
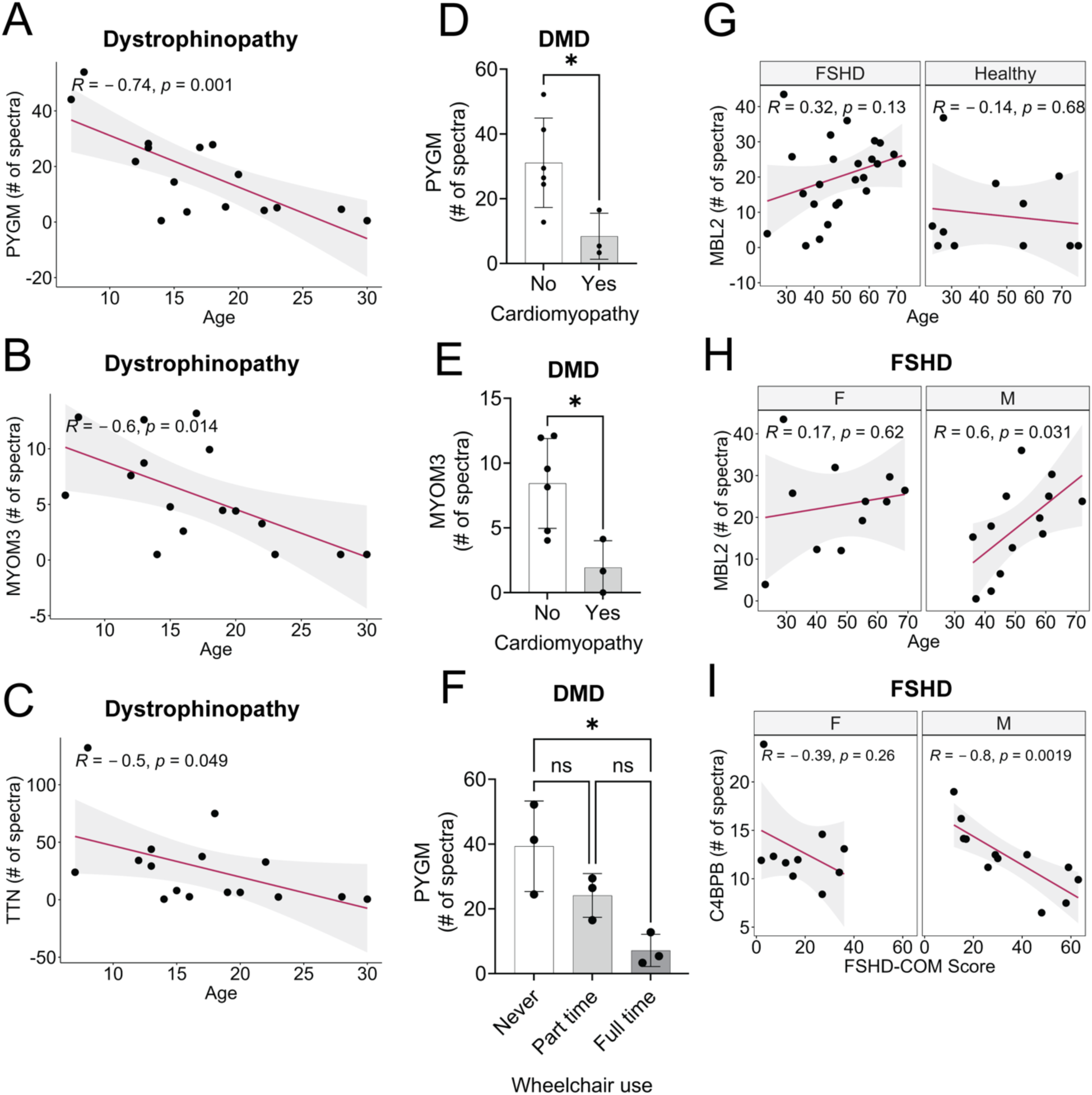
Correlations of EV-protein levels with clinical outcome measures and confounding factors. **(A-C)** Dystrophinopathy cohort (DMD and BMD). Pearson correlations of PYGM, MYOM3, and TTN with age. **(D-F)** DMD subset. PYGM and MYOM3 levels in patients with wheelchair use and cardiomyopathy. **(G-I)** FSHD cohort. Pearson correlations of MBL2 with age, and C4BPB with FSHD-COM score. The Pearson correlation coefficient (R) and statistical significance *p*val is computed for each EV-protein. Confidence intervals (95%) for each correlation are indicated by gray shade. ∗ *p*val < 0.05 obtained using two-tailed unpaired t test or one-way ANOVA.

We observed a trend of positive correlation of MBL2 levels with age in FSHD patients (*R*=0.32, *p*val: 0.13) (**Fig. 3F**). When sex is analyzed separately, we observed that this stemmed from the male cohort in which the MBL2 levels significantly increased with age (*R*=0.6, *p*val: 0.031) (**Fig. 3G**). It is noteworthy that, in the differential protein expression analyses (**Fig. 2C-D**), MBL2 was identified as a potential biomarker only in the female cohort. The correlation analyses suggest that while female patients maintain an overall elevated levels of MBL2 in all ages, MBL2 levels reach equivalent levels with age in male patients. Moreover, we observed a significant negative correlation in C4BPB levels in male FSHD patients with FSHD-COM score (*R*=-0.8, *p*val: 0.0019) (**Fig. 3H**). Accordingly, our results indicate that C4BPB is progressively decreased in patients with unfavorable FSHD-COM scores (i.e., higher scores). No significant correlation was observed for the other tested potential biomarkers and confounding factors (**Figs. S2-3**). Collectively, our analyses revealed the subset of potential biomarkers that correlate with either age (PYGM, MYOM3, and TTN in dystrophinopathy, MBL2 in FSHD) or FSHD-COM score (C4BPB in FSHD).

### Proteinase K treatment separates plasma proteins from EV proteins

Plasma proteome is mostly constituted of albumins, globulins, and fibrinogens. Moreover, certain plasma proteins, such as complement pathway components and lipoproteins, naturally form a corona layer around circulating EVs [43]. While SEC efficiently separates small nanoparticles from protein fractions, it is important to rule out any carryover plasma proteins in our analysis. Therefore, we treated EVs with 20 µg/mL of Proteinase K for 1h at 37°C, which selectively degrades proteins external to EVs. As Proteinase K cannot penetrate through the lipid bilayer of EVs, EV proteins are resistant to this treatment [44]. This treatment is followed by Proteinase K inhibition using 20 µM PMSF. This is essential to prevent Proteinase K from degrading EV intraluminal proteins upon their extraction from EV in preparation for immunoblotting (**Fig. 4A**). This experimental set up allowed us to eliminate the lipoprotein ApoB, while retaining the canonical EV marker CD9.

**Figure 4.**
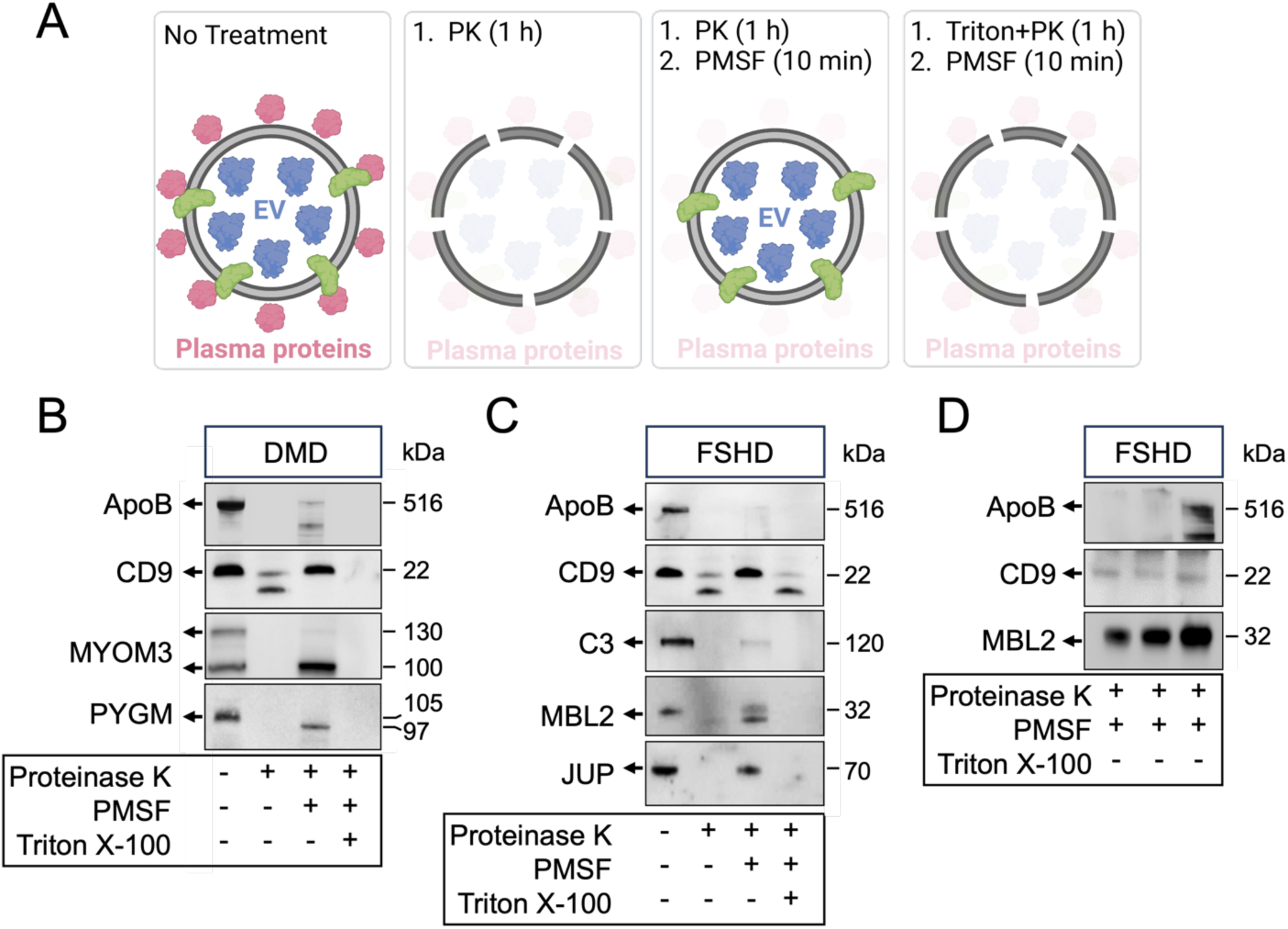
Confirmation of packaging or association of potential protein biomarkers within EVs. **(A)** Illustration of study design. EVs were treated with 20 µg/mL Proteinase K in the presence of 20 µM PMSF or 1% Triton X-100. Proteinase K resistant proteins (Lane 3) are considered EV proteins. **(B-C)** Displaying the abundance of potential biomarkers of Dystrophinopathy **(B)** and FSHD **(C)** using classical markers of plasma lipoprotein (ApoB), EV (CD9) and plasma complement pathway protein (C3) as controls. **(D)** Repetition of Proteinase K treatment of three new independent EV samples showing MBL2 is retained in EVs (uncropped blots are in **Supplementary fig. S5**).

Among the potential dystrophinopathy biomarkers, we observed that the smaller MYOM3 fragment is completely retained in EVs upon the full Proteinase K treatment, suggesting that it is packaged within EVs (**Fig. 4B**). Similarly, PYGM is also partially retained, suggesting that it is associated with EVs (**Fig. 4B**). In the FSHD cohort, we observed partial resistance of MBL2 and JUP to the full Proteinase K treatment, suggesting their association within EVs. MBL2 resistance to Proteinase K is particularly noteworthy as another complement component, C3, is completely degraded by the treatment (**Fig. 4C**), indicating a selective packaging of some elements of the complement pathway into EVs. We repeated the MBL2 western blot in three additional samples (**Fig. 4D**). Altogether, our results highlight the association of these biomarkers with plasma EVs and rule out unspecific contamination from plasma.

## Discussion

In clinical trials for DBMD and FSHD, outcome measures–such as clinical, imaging, biopsy-based and functional assessments–often lack the sensitivity to detect subtle changes in affected tissues. Additionally, they are invasive, costly, time-consuming, and may vary across sites, complicating patient stratification and delaying therapy approvals. Circulating molecular biomarkers offer a promising solution as non-invasive, cost-effective, objective and consistent surrogate endpoints. They can enhance therapy assessment, facilitate patient stratification, enable earlier detection of disease onset and improve long-term monitoring of disease progression across clinical sites. In this work, we set to explore the proteome of patient-derived plasma EVs to reveal disease-specific signatures. Thereby, we identified proteins exclusively and differentially expressed in dystrophinopathy and FSHD1 patients. Here, we focused on FSHD1 patients, as they represent the majority of FSHD cases. However, FSHD2 patients will be included in future studies due to the possibility of identifying a distinct EV-associated circulating biomarker profile, which could guide the appropriate selection and monitoring of FSHD2 patients in clinical trials. In this initial study, the differential EV cargo between DBMD and FSHD cohorts underscores the sensitivity of the assay and indicates that the observed proteomic signatures are specific to these distinct muscular dystrophies rather than generic inflammatory or muscle degenerative processes.

Under physiological conditions, the majority of released EVs is taken up by neighboring cells [45, 46]. Accordingly, the EV pool of the plasma is primarily originated from blood cells, with partial contributions of various organs and tissues [47]. However, upon injury or tissue damage, the local injured environment may favor more EV leakage to the circulation [48]. We believe that a muscular dystrophy would propel skeletal muscle derived EVs into circulation. While we cannot pinpoint the source of EVs in our plasma EV preparations, we can speculate that the skeletal muscle proteins identified in dystrophinopathy patients are derived from a pool of EVs released from damaged skeletal muscles. Dystrophin is an essential cytoskeletal stabilization protein in skeletal muscle. Upon the destruction of muscles in the absence of dystrophin, the structural muscle proteins (MYOM3, MYH2, FLNC, and TTN) may in part be packaged out of the damaged myofibers in EVs. Furthermore, the muscle associated glycogen phosphorylase PYGM has been reported to be packaged in EVs by cardiomyocytes upon cardiotoxin damage [49]. Another possibility is that one or more of these proteins are first released into blood as soluble factors and captured or encapsulated by EVs already present in the plasma. This is supported by the detection of TTN, which is known to be enriched in DMD biofluids, in our EV samples [50]. Notably, myofibrillar structural protein MYOM3 is also detected in sera of DMD and limb-girdle muscular dystrophy type 2D patients in two fragments of 100 kDa and 130 kDa [42]. While we were unable to perform necessary immunoblotting experiments on TTN due to its sheer size, we illustrated that the 100 kDa MYOM3 and PYGM are intraluminal EV proteins.

As we correlated the levels of these proteins to disease confounding factors, we identified that the levels of PYGM and MYOM3 are significantly higher in younger dystrophinopathy patients and are reduced in DBMD patients who have a history of cardiomyopathy. Furthermore, PYGM is significantly reduced in patients that are full time wheelchair users. Altogether, these results suggest that the EV-associated MYOM3 and PYGM may peak in early disease development and steadily decline as disease progresses. While we found no correlation of biomarker levels with steroid use, extending these correlations with newly approved dystrophin restoration therapies will be invaluable, as it could facilitate the validation of these biomarkers as surrogate endpoints.

On the other hand, no specific skeletal muscle related signatures are detected in FSHD patients where the skeletal muscle destruction is not as pronounced as observed in dystrophinopathies. Plasma EVs are very prone to acquiring apolipoproteins, fibrinogens, complement proteins, and immunoglobulins circulating in crude plasma [43, 51, 52]. These plasma proteins primarily form the corona layer of EVs. Originally speculated to be co-precipitations in EV preparations, these proteins are now established as core components of plasma EVs which provide the structural integrity to EVs and enhance their floatation density [43, 53]. In FSHD patients, we specifically observed differential expression of complement proteins in plasma EVs. Mild elevations of crude plasma complement components have been reported in FSHD patients. These included the <1.5-fold increases in C3 and C4b [23]. Our results support the notion of elevated complement activation in FSHD patients. Notably, in our EV samples a different set of complement proteins are identified. These include an increase in Complement C9 in the male cohort, a decrease in complement pathway inhibitor C4BPB in the male cohort, and an increase in mannose binding lectin (MBL2) in the female cohort (**Fig 2C, D, G and H**). Moreover, observed changes in these proteins are far more substantial than previously reported (MBL2 is upregulated 5-fold, C9 is upregulated 2.4-fold, and C4BPB is downregulated 2-fold). Thereby, it can be speculated that neglected changes in complement components can be unveiled by profiling these proteins that are either attached to or packaged within EVs. In particular, we provided evidence that MBL2 is at least partially packaged within EVs, extending the role of complement proteins beyond corona formation. This is in line with the previous reports on astrocyte derived EVs packaging MBL2 in EVs upon traumatic head injury [54]. While MBL2 is exclusively enriched in the female cohort, we also observed that in male FSHD patients its levels progressively increased with age. Whether MBL2 contributes to gender specific differences in FSHD needs to be tested in future studies.

Another key observation from the FSHD cohort was the exclusive expression of desmosomal proteins JUP and DSP in male patients. Interestingly, JUP has previously been reported to be upregulated in skeletal muscles overexpressing DUX4 [55], while an increase in DSP was observed in patient muscle biopsies [56]. In future studies, EVs released from FSHD cell lines can be collected and assayed for these two proteins to correlate their levels with DUX4 expression. Given the imbalanced sample size in the current male cohort (13 FSHD patients vs. 3 healthy controls), additional validation through an independent cohort with a more balanced patient-control ratio is warranted to fortify the observed findings.

Finally, we observed a decrease in GPLD1 in female FSHD patients. There is accumulating evidence that the expression of GPLD1 is regulated by insulin. Serum GPLD1 is in turn involved with adipokine Glypican-4 function, which normally interacts with insulin receptor on the cell membrane [57]. Whether the decrease in EV-associated GPLD1 has any impact in energy metabolism in female patients needs to be investigated.

Overall, our EV profiling reveals disease specific signatures found in circulation in DBMD and FSHD1 patients. It also provides key correlations with outcome measures and disease cofactors that highlight their potential use as clinical molecular biomarkers. The variable natures of muscular dystrophies such as DBMD and FSHD necessitate validation of our results in larger independent cohorts. Our current analysis serves as a primer which the follow up validations and longitudinal analyses could be built upon.

## Data Availability

All data produced in the present work are contained in the manuscript.

## Acknowledgments

We would like to thank Sarah Atkins, Madison Schmelzer, and Leigh Gabel for assistance in obtaining plasma samples, and to thank study subjects for their participation. This work was supported by S.Q.H. Nationwide Children’s Hospital retention funds, and a grant from the Chris Carrino Foundation for FSHD to N.Y.S. Also, portions of this work were supported by grant from the NIH/*National Institute of Arthritis and Musculoskeletal and Skin* Diseases Center of Research Translation in Muscular Dystrophy Therapeutic Development (P50AR070604) to K.M.F. and from the NIH/Eunice Kennedy Shriver National Institute of Child Health and Human Development (P50HD117373) to K.M.F and N.Y.S.

## Author Contributions

M.B.B., and N.Y.S. contributed to the conception and design of the study; M.B.B., D.H., K.T.N., and R.T. contributed to the acquisition and analysis of the data; M.B.B., E.R., K.M.F., S.Q.H., and N.Y.S. contributed to drafting the text and/or preparing the figures.

