## Supplemental Figures for "Identification of disease-specific extracellular vesicle-associated plasma protein biomarkers for Duchenne Muscular Dystrophy and Facioscapulohumeral Muscular Dystrophy"

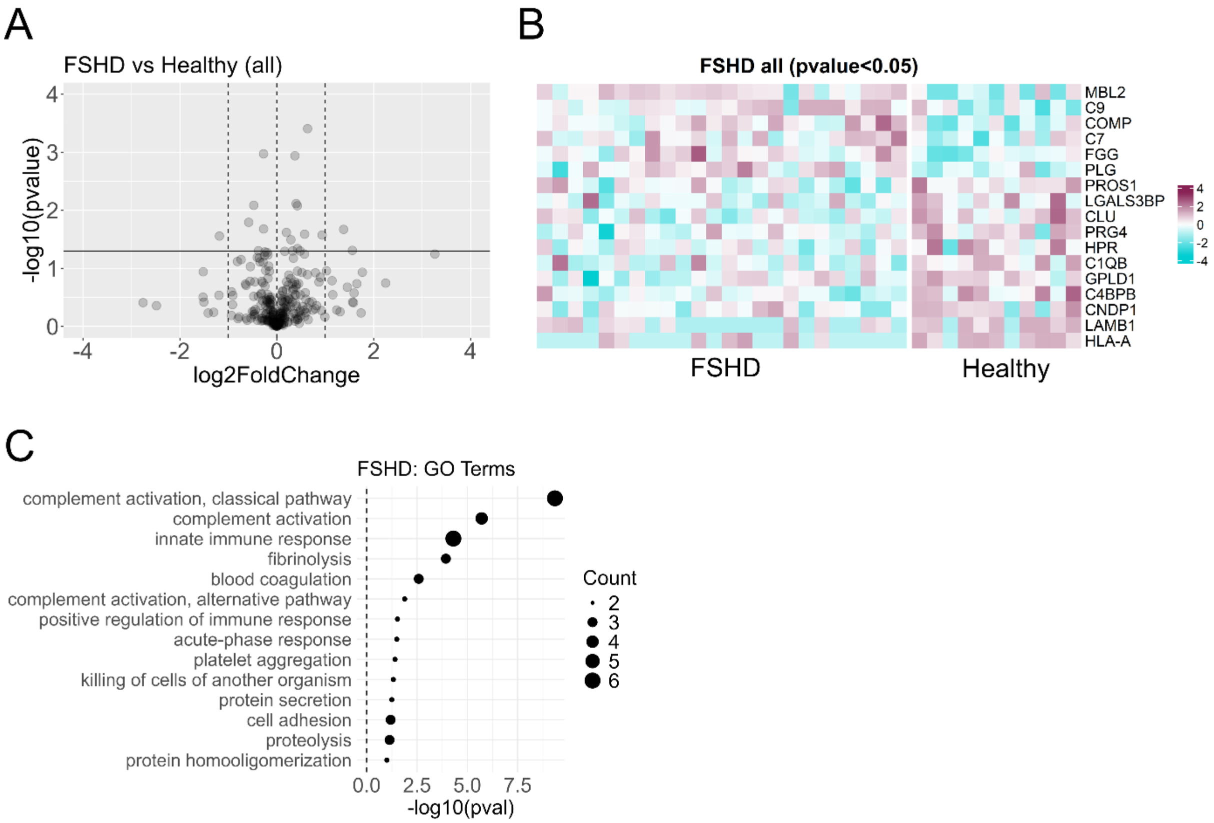


**Figure S1.** Differential protein levels in plasma EVs of FSHD1 patients and healthy controls. (**A**) Volcano plot of the FSHD1 cohorts combining both male and female patients. **(B)** Heatmaps of relative levels (i.e., z-scores) displaying all proteins that are significantly changing using nominal *p*val<0.05 (**C**) Enrichment analysis on the FSHD1 cohort conducted with DAVID software for Gene Ontology (GO) Terms with Biological Processes as sub-analysis. All significantly enriched terms are displayed (Bonferroni corrected pval<0.05).

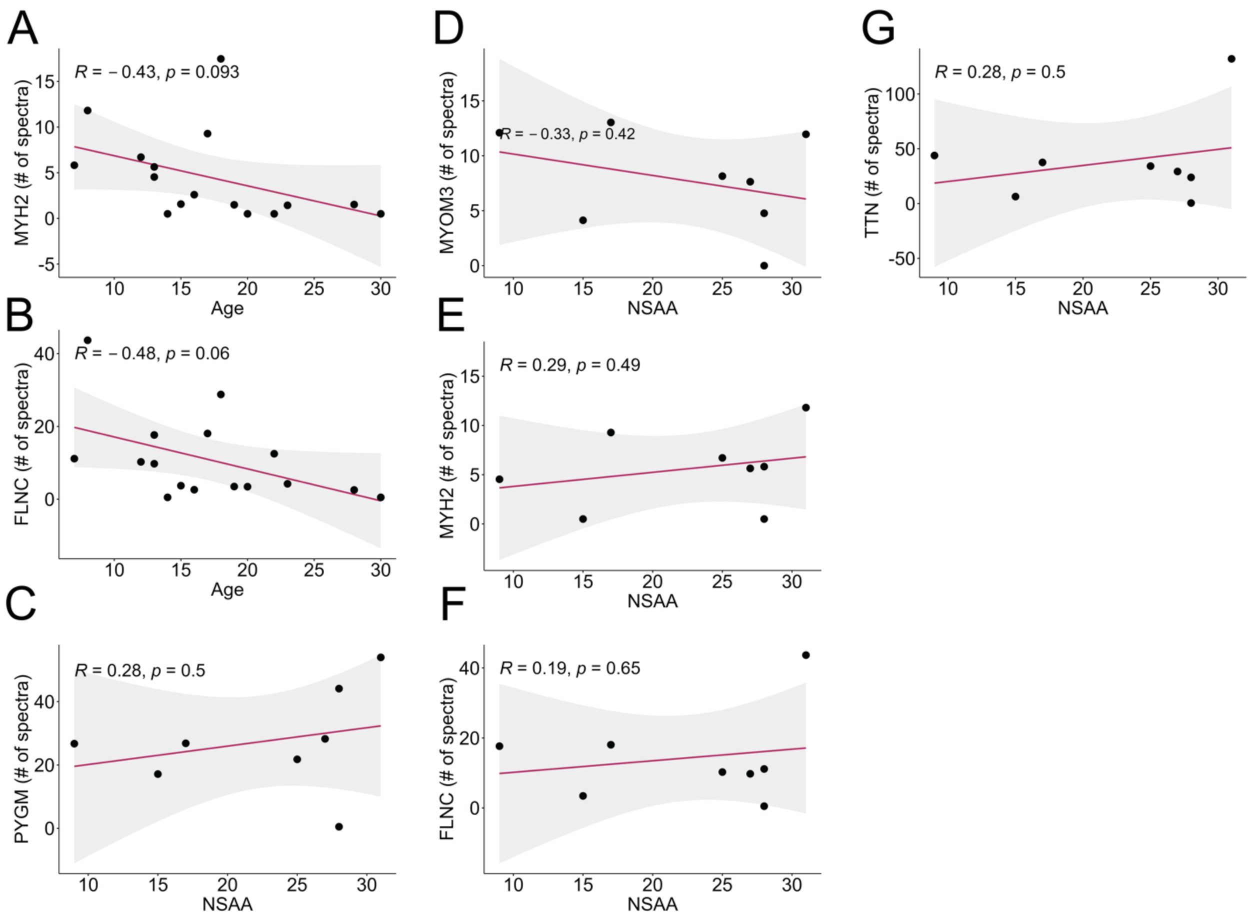


**Figure S2.** Correlations of EV-protein levels with clinical outcome measures and confounding factors in the dystrophinopathy cohort (DMD and BMD). **(A-C)** Pearson correlations of MYH2 and FLNC with age. **(D-H)** Pearson correlations of PYGM, MYOM3, MYH2, FLNC, and TTN with North Star Ambulatory Assessment (NSAA). The Pearson correlation coefficient (R) and statistical significance *p*val is computed for each EV-protein. Confidence intervals (95%) for each correlation are indicated by gray shade.

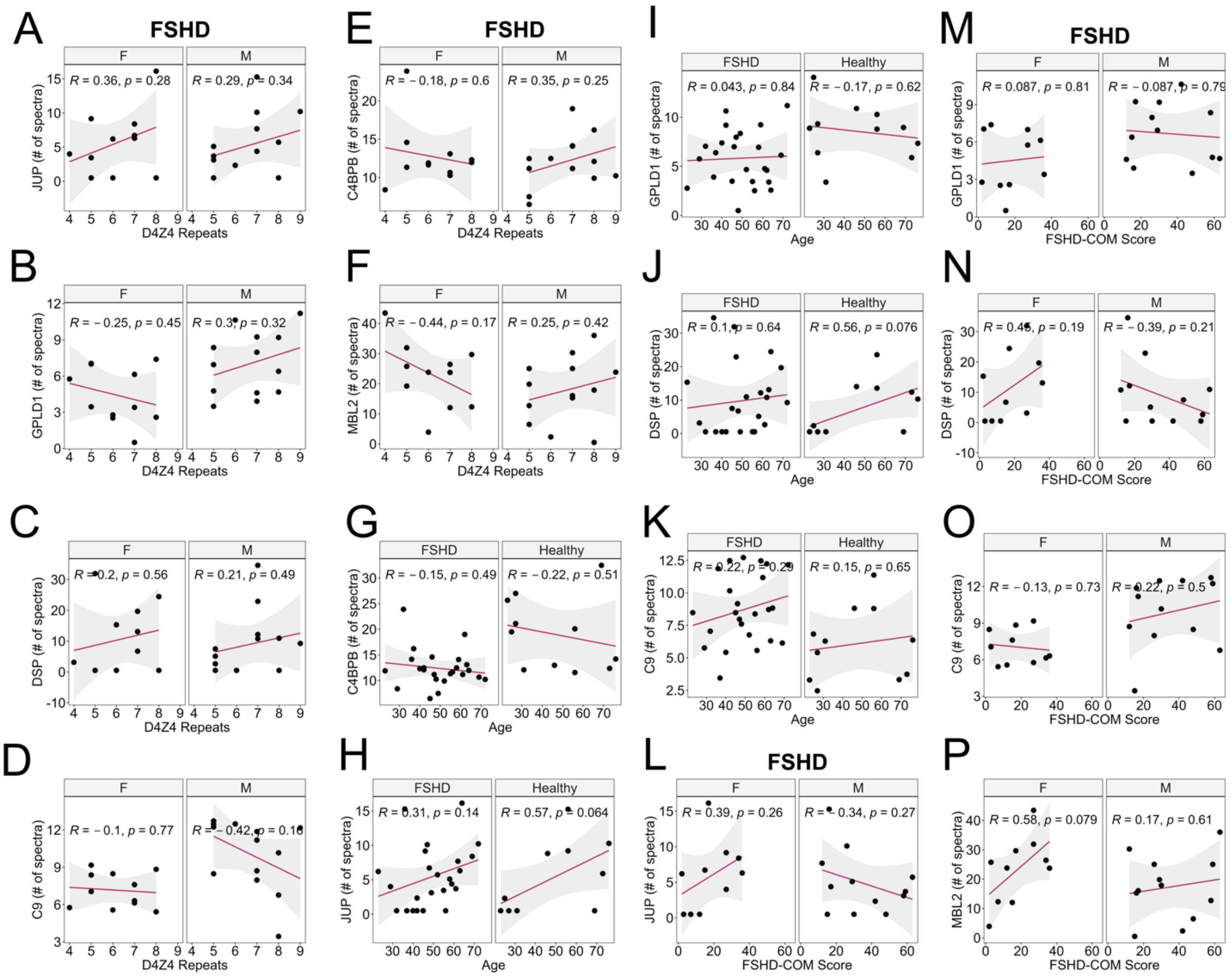


**Figure S3.** Correlations of EV-protein levels with clinical outcome measures and confounding factors in the FSHD cohort. **(A-F)** Pearson correlations of JUP, GPLD1, DSP, C9, C4BPB, and MBL2 with D4Z4 repeat length. **(G-K)** Pearson correlations of C4BPB, JUP, GPLD1, DSP, and C9 with age in female (F) and male (M) FSHD patients. **(L-P)** Pearson correlations of JUP, GPLD1, DSP, C9, and MBL2 with FSHD-COM score in female (F) and male (M) FSHD patients. The Pearson correlation coefficient (R) and statistical significance *p*val is computed for each EV-protein. Confidence intervals (95%) for each correlation are indicated by gray shade.

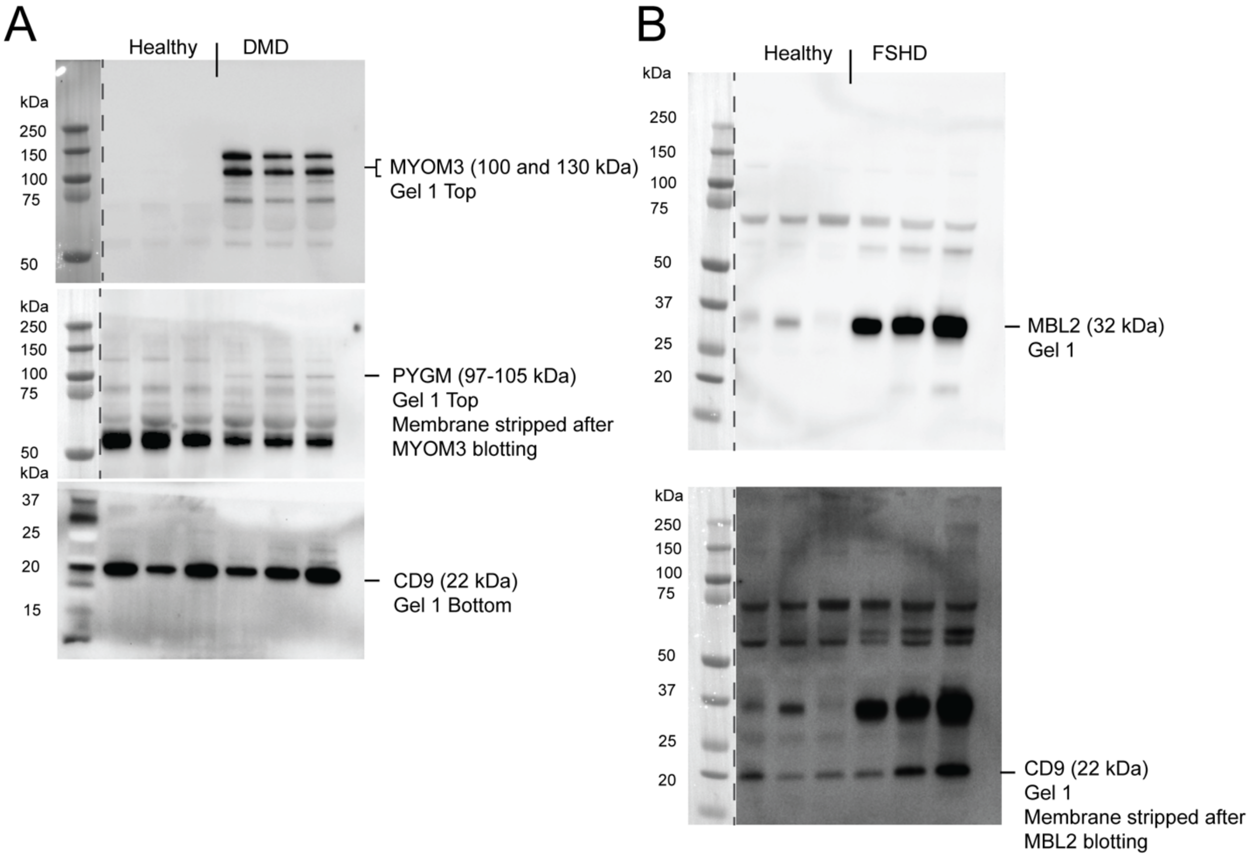


**Figure S4.** Uncropped western blots for figure 2J-K.

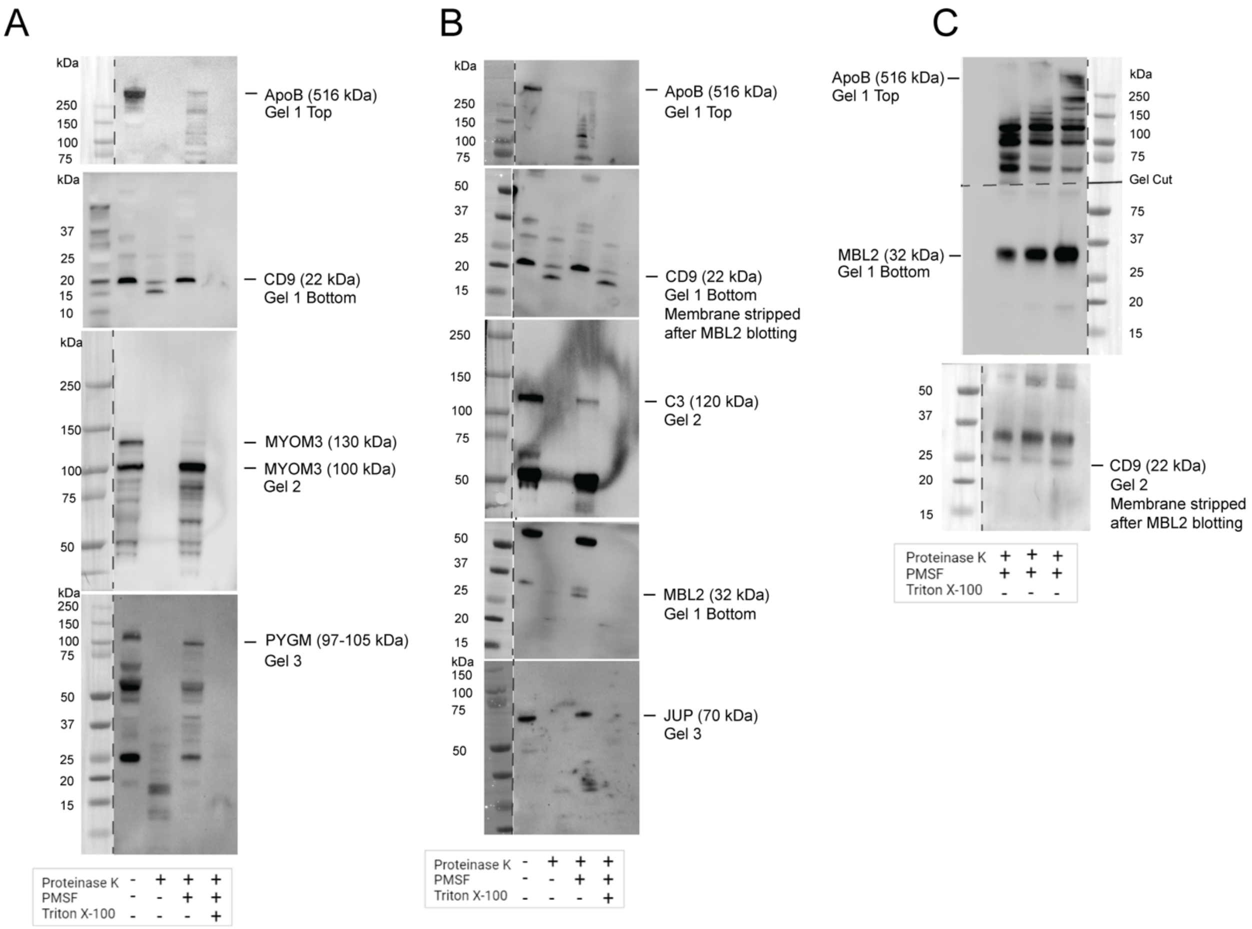


**Figure S5.** Uncropped western blots for figure 4B-D.
